# CERC-002, a human anti-LIGHT mAb reduces respiratory failure and death in hospitalized COVID-19 ARDS patients

**DOI:** 10.1101/2021.04.03.21254748

**Authors:** David S. Perlin, Garry A. Neil, Colleen Anderson, Inbal Zafir-Lavie, Lori Roadcap, Shane Raines, Carl F. Ware, H. Jeffrey Wilkins

## Abstract

**Background:** Severe COVID-19 infection is associated with dysregulated immune response which, in a substantial minority of patients, results in cytokine release syndrome (CRS) and acute respiratory distress syndrome (ARDS). Inhibition of cytokines or cytokine-associated signal transduction is a promising strategy to ameliorate ARDS associated with CRS. We and others have previously shown that serum free LIGHT (TNFSF14) levels are markedly elevated in patients with COVID-19 pneumonia/ARDS^10,11^, suggesting that LIGHT neutralization may offer therapeutic benefit to COVID-19 ARDS patients.

**Methods:** We conducted a randomized, double-blind, placebo-controlled, multi-center, proof-of-concept clinical trial of CERC-002 in adults with mild to moderate ARDS associated with COVID-19 (n=83). Enrolled patients received a single dose of CERC-002 or placebo, in addition to standard of care that included high dose corticosteroids. The primary efficacy endpoint was alive and free of respiratory failure status through Day 28. Secondary outcomes included alive status at Day 28, free of invasive ventilation through Day 28, and serum free LIGHT levels.

**Results:** In patients hospitalized with COVID-19 associated pneumonia and mild to moderate (ARDS), CERC-002 increased the rate of alive and free of respiratory failure status through Day 28 as compared to placebo (83.9% vs 64.5%; p=0.044). Efficacy was highest in the prespecified subgroup of patients 60 years old and older (76.5% vs 47.1%; p=0.042), the population most vulnerable to severe complications and death with COVID-19 infection. Through both the initial 28-day and 60-day follow-up periods, reductions of approximately 50% in mortality were observed for CERC-002 compared to placebo (7.7% vs 14.3% at Day 28 and 10.8% vs 22.5% at Day 60). Importantly, this improvement was incremental to standard of care including high dose steroids and remdesivir 88.0% and 57.8%, respectively). In addition, serum LIGHT levels but not IL-6 levels were markedly reduced in patients treated with CERC-002.

**Conclusions:** The data presented herein demonstrate that CERC-002 markedly reduces the risk of respiratory failure and death incremental to standard of care including high dose corticosteroids and reduces LIGHT levels in patients with COVID-19 ARDS. (ClinicalTrials.gov number NCT04412057).

## Introduction

Corona Virus Disease 2019 (COVID19) became global pandemic in March 2020. Ever since, millions of people have been infected with the SARS-CoV2 virus around the world, and the disease has and still killing many thousands each day. Most SARS-CoV2 patients have no or mild to moderate symptoms; however, many, particularly the elderly and those with concurrent health conditions, may have severe and life-threatening disease, including acute respiratory distress syndrome (ARDS) and cytokine release syndrome (CRS). The latter is thought to be major cause of severe morbidity and mortality^1 2,3^. While considerable progress has been made in controlling the epidemic with the introduction of vaccines^4^ and other novel therapies^5^, the appearance of SARS-CoV2 variants such as B.1.1.7 ^6,7^ and 501.V2^3^ presenting higher affinity to ACE2, appear to have increased infectivity and higher spread rates in the population^8^. Concerns have arisen that existing vaccines and therapies may be less effective in preventing and treating these variants^9,10^. Thus, there is an urgent need for drugs that mitigate severe illness in severely ill patients with CRS.

A number of cytokine neutralizing strategies have been studied. For example, IL-6 receptor blocker sarilumab^11^, IL-6 neutralization by tocilizumab^12,13^ have demonstrated limited efficacy.

LIGHT (also known as TNFSF14) belongs to the tumor necrosis factor superfamily and functions as an activator of both innate and adaptive immune responses. LIGHT belongs to a network of cytokines and receptors that create a self-regulating, host defense system^14^, and has a key role in the communication system which controls immune response. More specifically, LIGHT mediates immune activation and tissue damage. We have recently reported high levels of free (active) LIGHT in serum of COVID-19 patients and that these higher levels correlate with severity of disease^15^. These results were independently confirmed by Arunachalam et al^16^ and others ^17,18^. We therefore sought to assess the effectiveness of the human LIGHT neutralizing antibody, CERC-002, as a treatment for CRS and ARDS in COVID-19 patients.

Here, we report results of a randomized, double-blind, placebo controlled, multi-center, phase 2 clinical trial, which demonstrates a successful proof of concept for LIGHT neutralizing antibody, CERC-002, in patients hospitalized with mild to moderate COVID-19 ARDS.

## Methods

### Design

The trial was designated ClinicalTrials.gov number NCT04412057. Inclusion criteria for enrollment were as follows: Subject was ≥18 years of age at the time of informed consent and assent (as applicable); subject was male or non-pregnant, non-lactating female; subject had a diagnosis of COVID-19 infection through an approved testing method; subject had been hospitalized due to clinical diagnosis of pneumonia with acute lung injury defined as diffuse bilateral radiographic infiltrates and mild to moderate ARDS defined as PaO_2_/FiO_2_ >100 and <300 subject had an oxygen saturation at rest in ambient air <93%. Subjects were excluded if they were intubated with mechanical ventilation, currently taking immunomodulators or anti-rejection drugs), had received an immunomodulating biologic drug within 60 days of baseline, were in septic shock defined as persistent hypotension requiring vasopressors to maintain mean arterial pressure (MAP) of 65 mm Hg or higher and a serum lactate level greater than 2 mmol/L (18 mg/dL) despite adequate volume resuscitation, or had received any live attenuated vaccine, such as varicella-zoster, oral polio, or rubella, within 3 months prior to the baseline visit.

The primary endpoint was defined as the proportion of subjects alive and free of respiratory failure including one of the following: endotracheal intubation and mechanical ventilation, oxygen delivered by high-flow nasal cannula (heated, humidified, oxygen delivered via reinforced nasal cannula at flow rates >20L/min with fraction of delivered oxygen ≥0.5), noninvasive positive pressure ventilation, extracorporeal membrane oxygenation (ECMO).

Secondary endpoints were defined as mortality defined as the proportion of subjects who are alive at the Day 28/early termination (ET), partial pressure of arterial oxygen/percentage of inspired oxygen (PaO2/FiO2) ratio, time to invasive ventilation, duration of ventilatory support, duration of intensive care unit (ICU) stay, hospital length of stay, time to return to room air with resting pulse oximeter >93%, peak PaO2/FiO2 ratio, partial pressure of oxygen (PO2), change in sequential organ failure assessment (SOFA) score, change in body temperature, adverse event (AE) monitoring, and safety laboratory determination.

Exploratory endpoints were defined as viral load in nasopharyngeal aspirates, serum free LIGHT levels and inflammatory biomarker patterns, plasma concentrations of CERC-002 over time and measurement of anti-drug antibody (ADA).

Eligible patients were randomly assigned in 1:1 ratio to receive either CERC-002 or placebo. There were 11 trial sites in the United States. Enrollment began in July 2020 and ended in December 2020. A total of 82 patients were planned for randomization into the trial. This sample size was selected to provide greater than 80% power to detect a difference of 0.25 in the proportion of subjects alive and free of respiratory failure using a one-sided significance level of 0.05. This calculation assumed that the proportion alive and free of respiratory failure would be 0.60 in the placebo group and 0.85 in the CERC-002 group.

### Measurement of Serum Cytokines Levels

Free LIGHT assays were performed by Myriad RBM (Austin, TX, USA) using Quanterix’s fully automated HD-1 Analyzer and single molecule array (Simoa) technology as described elsewhere^19,20^. All incubations were performed at room temperature inside the Simoa HD-1 analyzer. Capture antibody conjugated paramagnetic beads were incubated with standards, samples or controls and biotinylated detection antibodies. The beads were then washed and incubated with streptavidin-ß-galactosidase (SßG). After the final wash, the beads were loaded into the Simoa Disc with enzyme substrate, resorufin ß-galactopyranoside (RGP). The fluorescence signals are compared to the standard curve and the quantity of free LIGHT was determined for each sample. The lower and upper limits of detection for free LIGHT were determined to be 0.8 to 4,000 pg/ml. IL-6 serum levels measured as a part of Myriad RBM InflammationMAP™ assay, on a Luminex platform, according to manufacturer protocol.

### Safety

The safety of CERC-002 was determined by the reporting of AEs, according to Common Terminology Criteria for Adverse Events (CTCAE) criteria (v 5.0), findings on physical exam, and the results of electrocardiograms (ECGs) and clinical laboratory tests.

### Study Objectives and Outcomes

The primary objectives were to evaluate the effect of CERC-002 compared with placebo, in addition to standard of care, in treating ARDS in adults which were diagnosed to have SARS-CoV-2 infection related pneumonia (COVID-19 pneumonia) and acute lung injury. The secondary objectives of the trial were to evaluate safety and tolerability of CERC-002 compared with placebo in addition to standard of care and to evaluate the effect of CERC-002 compared with placebo in addition to standard of care on mortality in adults with COVID-19 pneumonia and acute lung injury.

Exploratory objectives were to evaluate the effect of CERC-002 compared with placebo in addition to standard of care, on viral load in adults with COVID-19 pneumonia and acute lung injury, and to evaluate the pharmacokinetics (PK) and pharmacodynamics (PD) of CERC-002 in adults with COVID-19 pneumonia and acute lung injury.

### Statistical Analysis

All efficacy and safety variables were summarized using descriptive statistics. Descriptive statistics for continuous data included number of subjects (n), mean, standard deviation, median, minimum, and maximum. Summaries of change from baseline variables included only subjects who had both a baseline value and corresponding value at the timepoint of interest. Descriptive statistics for categorical data included frequency and percentage. The primary endpoint was alive and free of respiratory failure status through Day 28. Respiratory failure types were recorded as “Endotracheal Intubation and Mechanical Ventilation”, “Oxygen via High Flow Nasal Cannula”, “Noninvasive Positive Pressure Ventilation”, or “Extracorporeal Membrane Oxygenation (ECMO)”. For the purposes of the primary endpoint, use of respiratory failure was defined as either (a) having any new onset respiratory failure after administration of study drug at any time through Day 28 or (b) progression to endotracheal intubation and mechanical ventilation or ECMO at any time through Day 28 for those patients receiving non-invasive ventilation at the time of dosing. The proportion of subjects alive and free of respiratory failure through Day 28 in the CERC-002 group was compared to that in the placebo group using logistic regression methods. The logistic regression model included a fixed effect for treatment group. Model based point estimate (i.e., odds ratio), 90% confidence interval, and one-sided p-value were reported.

AE verbatim terms were coded using the Medical Dictionary for Regulatory Activities (MedDRA). The overall incidence of subjects having at least one AE were summarized. The incidence of treatment-emergent adverse events (TEAEs) was summarized by treatment group, system organ class (SOC), and preferred term (PT). Each subject was counted only once per SOC and PT. For continuous safety variables (e.g., laboratory and ECG measures), descriptive statistics for all reported and change from baseline values were summarized by treatment group and time point.

## Results

### Patients

Eligible patients were defined as hospitalized patients with documented COVID-19 Infection and clinical evidence of pneumonia with mild to moderate ARDS. As shown in Figure 1, 83 patients were deemed eligible to participate in the trial and were randomized either CERC-002 (41 subjects) or placebo (42 subjects). One randomized patient withdrew informed consent prior to receiving study drug and was not included in the study. Out of the 82 patients administered study drug, 62 patients had either no respiratory failure events prior to study drug administration or had a progression in their respiratory failure treatment; these 62 patients contributed to the intent to treat (ITT) analysis of the primary endpoint. All 82 patients administered study drug contributed to secondary efficacy analyses and safety analyses. The mean age of the CERC-002 group was 59.2, with 48.8% of patients above the age of 60 and 58.1 for the placebo group, with 50% of patients above the age of 60. Enrolled patients were given CERC-002 at dose of 16 mg/kg, with maximum dose of 1200 mg or placebo on day 1 by subcutaneous injection in addition to standard of care at the site. Patient disposition is shown in Figure 1.

**Figure 1:**
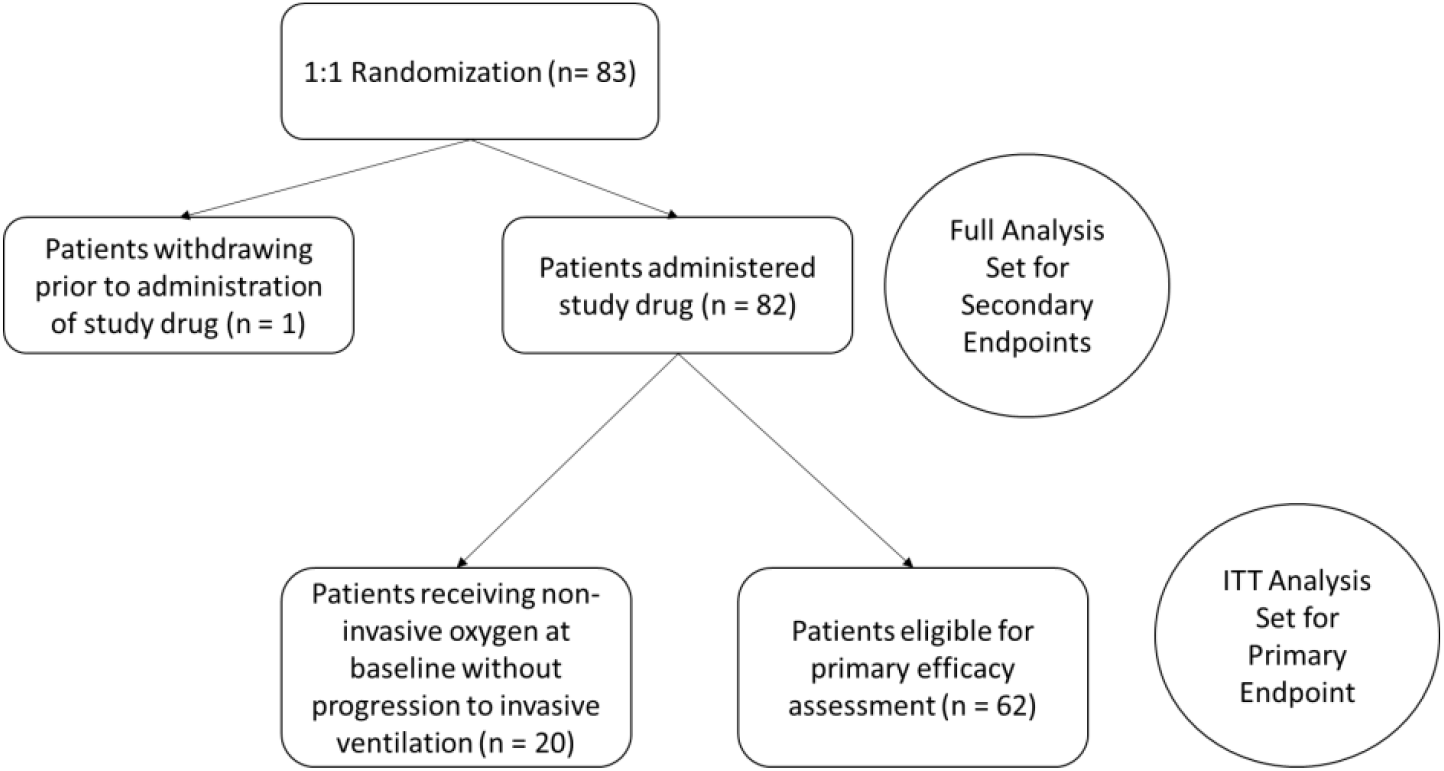
Enrollment and randomization

### Outcome

As noted, the primary endpoint was defined as the proportion of patients treated with CERC-002 compared with placebo in addition to standard of care, alive and free of respiratory failure through 28 days. As shown in Fig 2, patients treated with CERC-002 demonstrated a statistically significant improvement in in the rate of alive and free of respiratory failure in patients hospitalized with COVID-19 associated pneumonia, and mild to moderate ARDS (p=0.044, n=62). It is noteworthy that this reduction was incremental to standard of care, which included high dose steroids 88.0% and 57.8%, respectively of patients in the trial.

**Figure 2:**
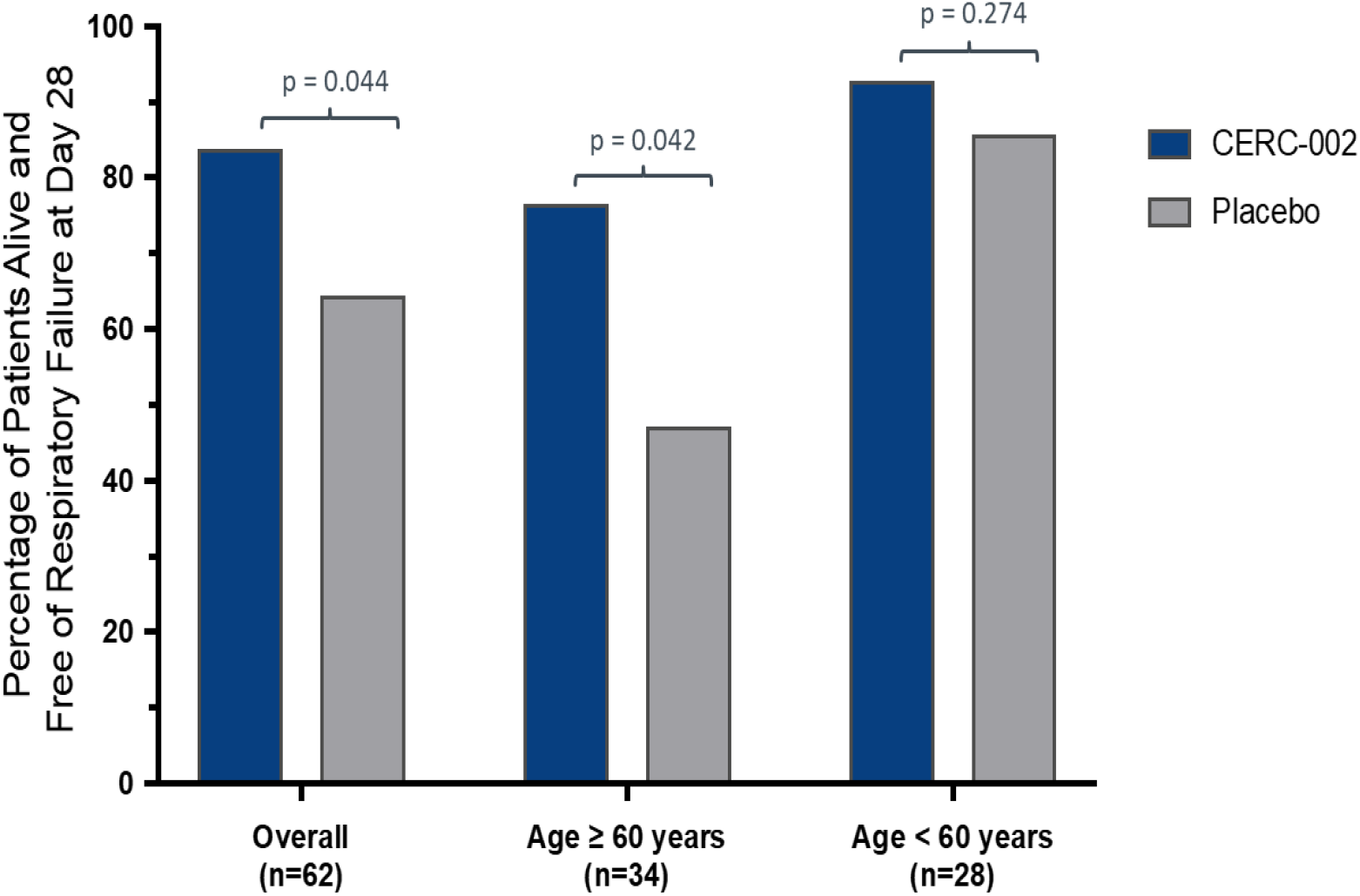
Efficacy of CERC-002 in COVID-19 patients. Percentage of patients alive and free of respiratory failure through 28 days after treatment is presented. Analysis was performed for overall (n=62) patients, and separately for subgroups of patients under the age of 60 (n=34) and age 60 or above (n=28).

A prespecified analysis showed that the benefit of CERC-002 was highest in patients 60 years of age and older (Fig. 2). Although not statistically significant, an approximately 50% reduction in mortality was observed in patients treated with CERC-002. This effect was maintained at the 60-day safety follow-up (Table 1).

**Table 1:** Improvements in mortality rate with CERC-002. Percentage of patients with death outcomes through both 28 and 60 days post-treatment.

|  | CERC-002 | Placebo |
| --- | --- | --- |
| **28-day Mortality** | **7.7%** | **14.3%** |
| **60-day Mortality** | **10.8%** | **22.5%** |

### Key Secondary outcomes

Free LIGHT levels were measured prior to administration of study drug on Day 1 and on several time points post treatment. As demonstrated in Figure ***3***, free LIGHT levels declined significantly (80%) and quickly in the CERC-002 treated group and increased somewhat in the placebo group (Figure *3*).

**Figure 3:**
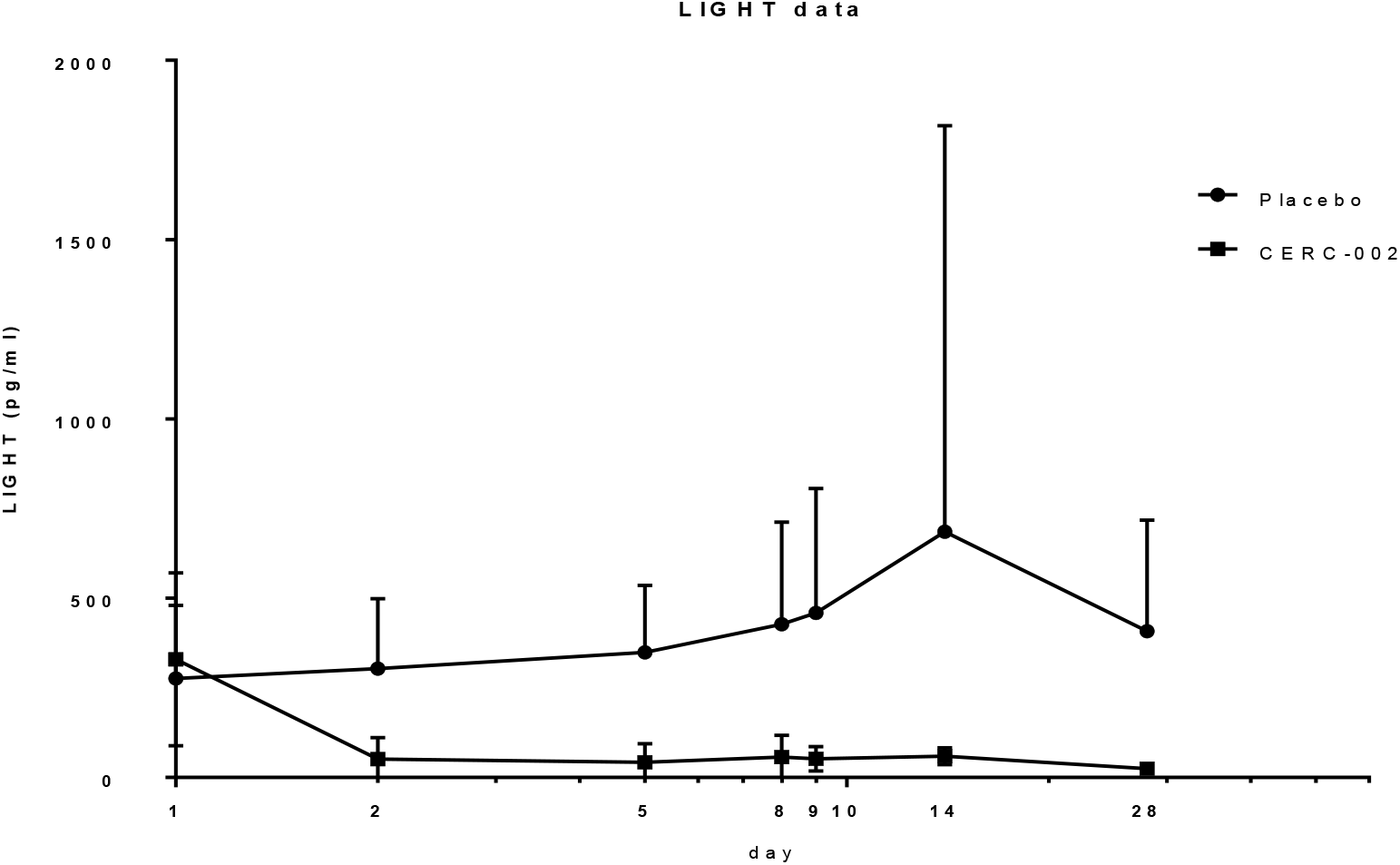
Free LIGHT Levels (pg/mL) over treatment period. Mean free LIGHT levels were comparable at baseline across cohorts.

### Safety and tolerability

CERC-002 was well tolerated as at a single dose of 16mg/Kg, with no serious adverse events attributed to CERC-002. Mild to moderate adverse events were detected in 16 patients (40%) on CERC-002. Of all adverse events, regardless of causality, leukocytosis (15%), Anemia 10%, hepatic enzyme increase (10%) acute kidney injury (7.5%) and respiratory failure (7.5%) as those adverse events with a frequency greater than 5% in this critically ill population. Comparison of adverse events across the two cohorts was comparable in adverse events and their frequency. There was no evidence of increased infections or adverse events related to immunosuppression in the CERC-002 cohort versus the placebo cohort.

## Discussion

This multicenter, randomized, double-blind, placebo-controlled, Phase 2 trial is the first demonstration of the use of LIGHT-neutralizing therapy as a treatment for hospitalized patients with COVID-19 pneumonia with ARDS. The results show substantial reduction in respiratory failure and death, incremental to steroids treatment and standard of care and a dramatic reduction in serum LIGHT. CERC-002 was well tolerated, and noteworthy for a lack of increased opportunistic infection and drug related serious adverse events.

COVID-19 patients who develop pulmonary symptoms such as acute lung injury and eventually ARDS demonstrate hyperactive immune properties, with the majority of COVID-19 ARDS patients demonstrating cytokine release syndrome^21^. A significant number of these patients succumb to the disease which is a major driver of the overall mortality associated with COVID-19.

Systemic corticosteroids have been shown to be effective in COVID-19 ARDS^22^, reducing mortality by about 30%^23^. Based on these results steroids have rapidly become standard of care in the treatment of severe COVID-19 ARDS. However, steroids have considerable liability, particularly in elderly patients, diabetic patients, and those with compromised immunity; in addition, steroids have not been studied in mild to moderate COVID-19 ARDS^24^. The effectiveness of steroids in this context supports the hypothesis that immunomodulation can benefit patients with severe COVID infection. In this study, we showed that CERC-002 showed clear benefit on top of steroid use without evident increased liability of immunocompromise. It is not clear from our results whether CERC-002 as a monothereapy would have provided similar benefit. This is clearly a topic for further study.

Several cytokine neutralizing strategies have been assessed for the treatment of COVID19 ARDS CRS patients. These include IL-6R or IL-6 cytokine neutralizing agents such as sarilumab^25^ and tocilizumab^12^. These agents show benefits on respiratory and cardiovascular organ support-free days endpoints but without clear benefit on mortality^26,27^. In our study, we also showed elevation in IL-6 levels in patients with COVID-19 ARDS. Interestingly neither steroid therapy nor CERC-002 showed a reduction in IL-6 levels. However, despite this, CERC-002 did show clear clinical benefit on top of corticosteroids, suggesting that LIGHT neutralization may have an independent effect on the course of this disease. Because CERC-002 does not appear to influence IL-6 levels, at least acutely, it is possible that combination LIGHT and IL-6 neutralizing therapy might have additional benefit. Again, this is a topic for further study in future trials.

The IL-1β neutralizing agent, Anakinra^28^ and additional IL-1β neutralizing agents have been assessed in clinical trials^29^. GM-CSF^30^ and VEGF^31^ neutralization were also assessed, with modest effect. Although initial results are promising, additional controlled studies are required to demonstrate conclusive efficacy.

LIGHT is well known for its multifaceted role in driving the immune system. From T-cells co-stimulation^32^, orchestrating fibrosis^33^ to driving unleashed immune response leading to autoimmunity^34^, LIGHT is a cytokine that influences many immune activating pathways. In light of the accumulating evidence of partial response to cytokine neutralizing agents and the understanding of COVID-19 immune profile, we have recently reported elevated free LIGHT levels in serum of COVID-19 ARDS patients, correlating with severity^15^. Moreover, the expression patterns of LIGHT receptor HVEM, in myeloid and in tissue-barrier epithelial cells indicates excessive LIGHT may cause accumulation of neutrophils, macrophages and T cells that promote tissue destruction^35^. This is further supported by evidence that LIGHT has a role in pulmonary fibrosis^36^. A major source of LIGHT are T-cells, macrophages and neutrophils^37^ which were reported to infiltrate the lungs during COVID-19 infection^38^. Moreover, LIGHT has a role in pulmonary inflammation driven by viral infection and its levels correlate with disease severity^39,40^. This evidence supports the involvement of LIGHT in COVID-19 related ARDS and cytokine release syndrome.

This study provides the first evidence that neutralizing LIGHT with a specific monoclonal antibody has potential utility and may be life-saving in COVID-19 related ARDS. Further, larger studies are needed to verify these data.

## Data Availability

The data that support the findings of this study are available from the corresponding author, [Garry A Neil], upon reasonable request.

## Supplementary

**Table S2:**
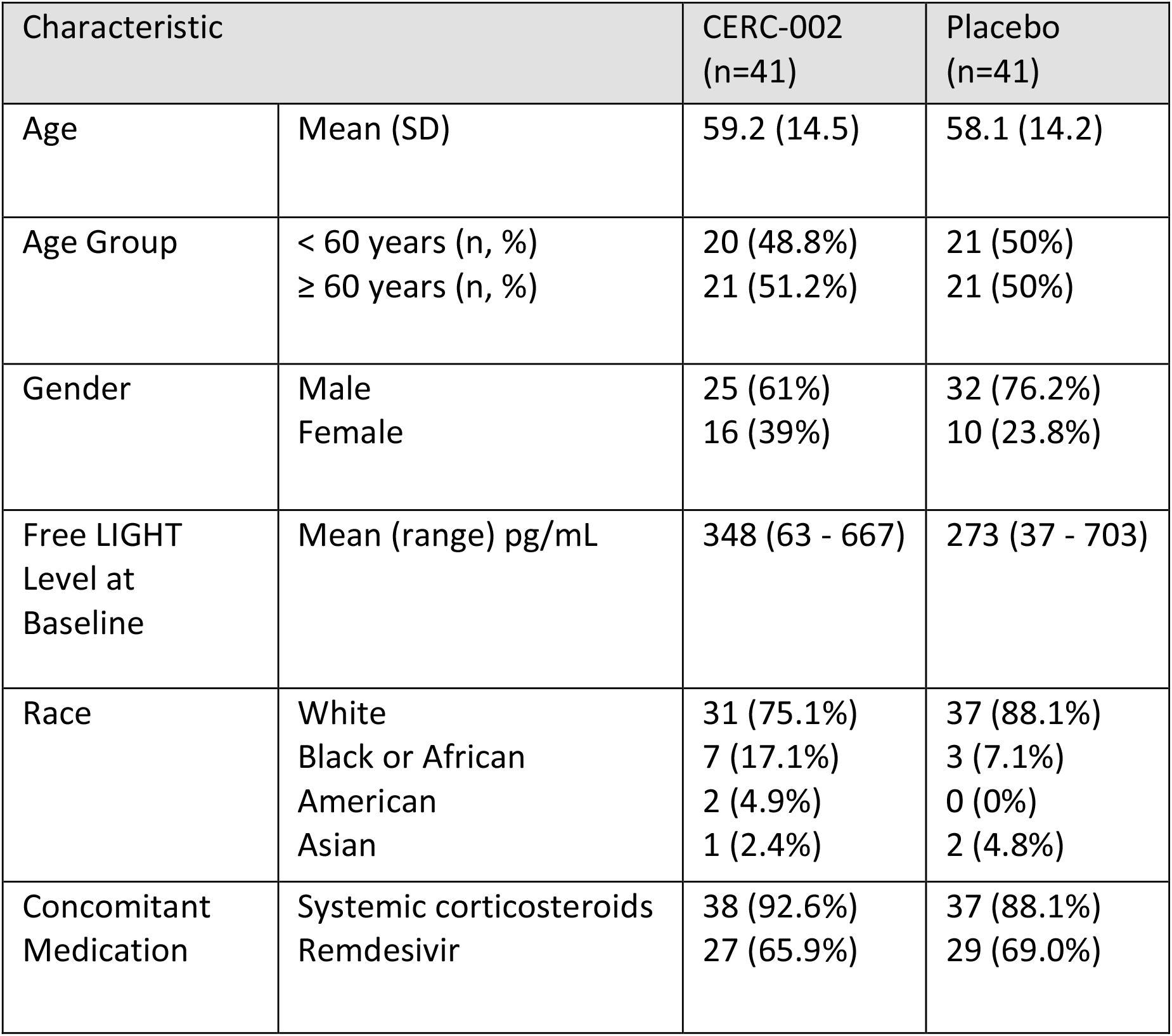
Patient Demographics

